# Clinical management of patients with multimorbidity: A qualitative exploration of healthcare worker perspectives in Malawi and Tanzania

**DOI:** 10.1101/2025.04.24.25326378

**Authors:** Ibrahim Gibunje Simiyu, Gift Treighcy Banda, Nateiya Mmeta Yongolo, Sangwani Salimu, Martha Oshosen, Grasiana Kimario, Alice H. Rutta, Jacob Phulusa, Gimbo Hyuha, Juma Mfinanga, Julian T. Hertz, Blandina T. Mmbaga, Sarah Urasa, Francis M. Sakita, Rhona Mijumbi, Charity Salima, Adamson S. Muula, Miriam Taegtmeyer, Jamie Rylance, Eve Worrall, Hendry Robert Sawe, Paul Dark, Felix Limbani, Ben Morton, Multilink consortium

## Abstract

**Background:** Multimorbidity, the coexistence of two or more chronic medical conditions, poses significant challenges for healthcare systems in sub-Saharan Africa (SSA), where single-disease-focused approaches currently predominate. Despite the rising burden of multimorbidity in SSA region, data on its clinical management in hospitals is limited. This study aimed to explore healthcare worker experiences in the management of patients with multimorbidity in Malawi and Tanzania.

**Methods:** We conducted in-person in-depth interviews from February 2023 to July 2023. We purposively selected healthcare workers from emergency, outpatient and internal medicine departments in Malawi and Tanzania. Our analysis utilized the Sustainable intEgrated chronic care modeLs for multi-morbidity: delivery, FInancing, and performancE (SELFIE) framework for integrated care for multimorbidity to categorize codes into corresponding domains and themes. The analysis examines workforce, service delivery, and finance domains, together with the core of the SELFIE framework (patient) factors and generated 11 sub-themes that influence clinical decision-making in these contexts.

**Results:** We interviewed 45 healthcare workers including clinicians, nurses and pharmacists involved in the management of patients with multimorbidity in district and tertiary hospitals. Healthcare workers noted that limited patient knowledge of chronic diseases; delayed hospital presentation; inadequate chronic disease training; and a lack of multimorbidity clinical practice guidelines limit high-quality clinical decisions. In addition, restricted access to diagnostics and medicines together with high out-of-pocket costs for chronic disease management further increase the challenge of multimorbidity management.

**Conclusion:** This study identified multiple domain factors that influence healthcare workers clinical decisions for the management of patients with multimorbidity revealing gaps in clinical training and clinical practice guidelines. Our findings emphasize the need to strengthen integrated care, expand workforce capacity, update clinical guidelines, and strengthen healthcare financing to address the growing multimorbidity burden in SSA.

## Introduction

Multimorbidity, the co-existence of two or more chronic medical conditions in the same individual at the same time (1), poses a major global health challenge to healthcare systems (2). Multimorbidity is associated with increased healthcare utilization and cost; reduced health-related quality of life; and increased risk of disability and mortality (2–4). The increased complexity of multimorbidity management is a major challenge for healthcare providers worldwide (5–8). This issue is particularly acute in sub-Saharan Africa (sSA) where life expectancy is increasing and there is a high prevalence of both chronic communicable and non-communicable diseases (9, 10); and where fragile health systems are frequently organised vertically, focussed on single diseases (11–13).

Studies conducted in high-income countries (HICs) demonstrate single disease-focused approaches for patients with multimorbidity may result in sub-optimal treatment and adverse outcomes (14, 15). These include increased treatment burden and polypharmacy (including increased risk of adverse drug reactions) (7); fragmented care delivery; and barriers to effective shared decision-making (6). There is a paucity of data from sSA settings on clinical care delivery, however, critical shortages of healthcare workers and resource constraints may further impede effective management for patients with multimorbidity (10, 16).

This study aims to explore healthcare worker experience of multimorbidity management in hospital settings in Malawi and Tanzania. It is vital to understand how healthcare providers conceptualize and manage multimorbidity to inform the development of context-sensitive health systems capable to address the evolving needs of the sSA population.

## Methods

This study was nested within Multilink, a prospective longitudinal cohort study which aims to establish the prevalence of multimorbidity among patients acutely admitted to hospitals in Malawi and Tanzania (17, 18).

### Study Design

We used a phenomenological approach to explore healthcare workers perspectives and experiences on the management of multimorbidity. Our adoption of this approach was to capture and describe the essence of participants subjective lived experiences on the management of patients with multimorbidity (19). We collected data through face-to-face in-depth interviews with healthcare workers in Malawi and Tanzania. This study is reported according to the Standard for Reporting Qualitative Research guidelines (SRQR) checklist (20) (see online supplementary appendix 1)

### Theoretical framework

We used the SELFIE (Sustainable IntEgrated Chronic Care modeLs for Multimorbidity: delivery Financing and performance) framework for integrated multimorbidity care to consider healthcare delivery according to the six WHO health systems building blocks (21). The SELFIE framework facilities dialogue by structuring integrated care concepts for multimorbidity and supports the description, development, and implementation of chronic care programs (22). The SELFIE framework also provides a comprehensive guide to evaluate and analyse multiple aspects of chronic disease management and care. The SELFIE framework is structured around the core, micro-, meso- and macro-levels (23). The micro-level involves interactions between individuals with multimorbidity and healthcare providers. The meso-level addresses the organizational structure of healthcare providers, while the macro-level encompasses legislation, governance, policies, and system-wide changes at national and international levels (23).

### Study sites and population

Whilst multimorbidity research has focused on primary and community settings in sSA (24), recent data highlight increasing burden in hospital settings (25, 26). Healthcare workers are commonly initially trained in hospital settings before progressing to independent practice in primary and community settings. The study was conducted in four public sector hospitals: the Queen Elizabeth Central Hospital and Chiradzulu District Hospital in Malawi; and Muhimbili National Hospital and Hai District Hospital in Tanzania. In Malawi, public healthcare services are provided free of charge (27), whereas in Tanzania, public healthcare operates on a cost-sharing and health insurance model (28). Queen Elizabeth Central Hospital is Malawi’s largest referral and tertiary teaching hospital with a bed capacity of 1350 located in Blantyre. Chiradzulu District Hospital is on the outskirts of Blantyre, serving the semi-urban population and has a bed capacity of 400. Muhimbili National Hospital is the national referral in Tanzania, with a bed capacity of 1500, located in Dar es Salaam. Hai District Hospital is in the Kilimanjaro region, serving the semi-urban population and has a bed capacity of 140 (29).

Study participants were healthcare workers from the four study sites who deliver care in emergency departments, outpatient departments or medical inpatient wards; recruited through purposive sampling and snowballing approaches (30). We selected participants to incorporate multidisciplinary team members (medical doctors, clinical officers, nurses and pharmacists) involved in the clinical management of patients with multimorbidity. Clinical officers are non-physician medical workers who support tasks previously performed by doctors; this cadre is present in the majority of SSA health systems (31, 32). Healthcare workers were eligible to participate in the study if: (1) they were actively working as healthcare providers in the emergency, outpatient or medical inpatient wards of the recruitment sites; (2) had at least three years of experience managing patients in hospital settings to ensure sufficient depth and breadth of perspectives on multimorbidity (33); and (3); provided written informed consent to participate.

### Data collection

Our interview guide was co-created to be generalisable across Tanzanian (led by IGS) and Malawian (led by GTB) study sites. This included questions to explore (1) healthcare worker understanding of multimorbidity; (2) clinical decision making for multimorbidity management; and (3) enablers and barriers to clinical decision making for multimorbidity management (see Supplementary appendix 2). The first two interviews with healthcare providers from each site were used as an internal pilot and iteratively develop interview guide; these interviews were incorporated within our analysis. Through this process it became clear that healthcare workers used the terms “comorbidity” and “multimorbidity” interchangeably in both Malawi and Tanzania, in line with previous literature (34). Therefore, we adopted a flexible terminological approach tailored to individual participant preference within the interviews.

Before data collection, FL led qualitative workshops to standardize data collection for researchers in Malawi and Tanzania and ensure a shared understanding of the interview guide and techniques. Interviews in Tanzania were conducted at Muhimbili National Hospital by IGS and GK; and at Hai District Hospital by MO. In Malawi, all interviews with healthcare workers were conducted by GTB. The interviews were conducted in Swahili (Tanzania), Chichewa (Malawi) or English (Tanzania and Malawi) within hospital premises for participant convenience; and lasted for approximately one hour. Given the heterogeneity of healthcare workers, we set a minimum sample of three participants per healthcare worker category at each site to ensure adequate representation (35). We conducted interviews until data saturation (no new information emerged from the participants) was achieved (36–38).

### Data management and analysis

Data was de-identified and transcripts assigned a unique ID number prior to upload on a secure server. All interviews were transcribed verbatim in the languages they were conducted and where necessary translated into English by IGS and MO in Tanzania and GTB in Malawi. Translated transcripts were exported into NVivo 12 Plus software (2021) (39) for coding and analysis. We applied the established framework method to manage and analyse the data (40). IGS read through all transcripts from Malawi and Tanzania and coded them separately in NVivo 12. The codes were deductively generated from the study objectives and inductively from the transcript through open coding. The initial codes were used to develop the coding framework that was reviewed by FL before finalisation. Codes were indexed and charted into the coding framework domains into themes and sub-themes (22, 41). Team discussions and meetings played a crucial role in clarifying and resolving differences in the understanding of the analytical framework. Our analysis explores the core (individual with multimorbidity); healthcare service delivery; healthcare workforce considerations; and healthcare finance domains of the SELFIE framework as these relate to the management of multimorbidity.

### Positionality

IGS is a qualified medical doctor in Tanzania and conducted this research study as part of his PhD focused on multimorbidity. IGS had a prior professional relationship with some of the participants interviewed in Tanzania but had no clinical responsibilities for the care of patients with multimorbidity during this study. GTB, a PhD fellow in the Multilink program with a background in physiotherapy and public health, and MO, a social scientist with public health training, used their expertise to engage effectively with healthcare workers. Piloting of the interview guide was therefore essential to underpin interpretation and exploration of questions posed to research participants. This approach facilitated a shared understanding of interview questions and approaches, enabling incorporation of multiple perspectives to enrich consistent questioning during the study.

### Ethical considerations

This study was approved prospectively by the Liverpool School of Tropical Medicine, United Kingdom (21-086; approved on 10.05.2022); the College of Medicine and Research and Ethics Committee, Malawi (P·11/21/3462; approved on 15.10.2021); Kilimanjaro Christian Medical College, Tanzania (2570; approved on 15.12.2022) and National Institute of Medical Research, Tanzania (NIMR/HQ/R·8a/Vol·IX/4008; approved on 13.05.2022). All participants provided written informed consent to participate in this study. Additionally, we made it clear to healthcare workers, that they could withdraw any time from the study without consequences, and their data would be removed from the study. Our reflexivity statement (appendix 3) describes how equitable research partnership and authorship between LMICs and HIC researchers was promoted within the Multilink consortium (42).

## Results

We interviewed 45 healthcare workers, 20 from Malawi and 25 from Tanzania, between February 2023 and July 2023. Out of the 45 participants, 63 % (25/45) were male and 37 % (20/45) were female (Table 1). The median duration of clinical practice experience was 8 years (range 3-34 years). Participants included a range of healthcare worker cadres, including specialist and generalist doctors, clinical officers, nurses and pharmacists (Table 1). We identified four broad themes that influence healthcare workers clinical decisions for the management of multimorbidity: 1) Individual patients with multimorbidity factors; 2) workforce; 3) service delivery and 4) financing (Table 2) and Appendix 4.

**Table 1:** Characteristics of healthcare providers interviewed in Malawi and Tanzania.

|  | Malawi (N=20)<br>n= (%) | Tanzania (N=25)<br>n= (%) | Total (N=45)<br>n= (%) |
| --- | --- | --- | --- |
| Gender |  |  |  |
| Male | 11 (55) | 14 (56) | 25 (63) |
| Female | 9 (45) | 11 (44) | 20 (37) |
| Type of facility |  |  |  |
| District hospital | 11 (55) | 10 (40) | 21 (47) |
| Tertiary | 9 (45) | 15 (60) | 24 (53) |
| Position |  |  |  |
| Specialist | 1 (5) | 6 (24) | 7 (16) |
| General medical doctor | 5 (25) | 4 (16) | 9 (20) |
| Clinical officer | 8 (40) | 1 (4) | 9 (20) |
| Pharmacist | 1 (5) | 2 (8) | 3 (7) |
| Nurse | 5 (25) | 12 (48) | 17 (37) |
| Experience (years) |  |  |  |
| 3-5 | 10 (50) | 4 (16) | 14 (31) |
| 6-10 | 4 (20) | 12 (48) | 16 (36) |
| 11+ | 6 (30) | 9 (36) | 15 (33) |

**Table 2:** Summary of themes and sub-themes.

| Domain | Level | Item | Sub-theme |
| --- | --- | --- | --- |
| Individual patient with multimorbidity | NA | Environment | Disease severity at hospital presentation<br>Literacy |
| Workforce | Micro | Multidisciplinary team | Delays in management<br>Absence of task shifting for multimorbidity |
|  | Meso | Continuous professional development | Lack of on-job training (post graduate training) |
|  | Macro | Education and workforce planning (Clinical education) | Absence of integrated disease management training |
|  |  | Workforce demography match | Misdiagnosis in lower-level facilities (Unmatched workforce distribution) |
| Service delivery | Micro | Treatment interactions | Lack of guidelines for multimorbidity |
|  | Meso | NA |  |
|  | Macro | Service availability and access | Investigations and service availability |
|  |  | Policies to integrate care across organizations and sectors | Lack of integrated policies |
| Financing | Micro | Out of pocket costs | Healthcare access and treatment costs |
|  | Meso | NA |  |
|  | Macro | NA |  |

### Individual patient with multimorbidity factors

#### Literacy/knowledge of chronic diseases

An understanding of chronic diseases among patients with multimorbidity is essential for effective management of their conditions. Healthcare workers in Malawi and Tanzania acknowledged that patient knowledge of chronic diseases influences clinical decision-making in multimorbidity management and emphasized the importance of patient health education.

> “*From my experience, most of the patients with chronic conditions lack the right information and knowledge about their conditions. They may be taking medications and stop when feeling better and end up with complications. We need to educate them at the point of diagnosis* ” [Specialist 4 Tanzania].

> “*There are patients that don’t know hypertension or diabetes, or CKD (chronic kidney diseases) are chronic diseases that require lifelong treatment. They will get their medication supply and when they feel better, they stop attending clinics* [General medical doctor 8 Malawi]

Healthcare workers raised concern about beliefs and use of alternative treatment among patients with chronic conditions from lack of information and limited health literacy. Some healthcare workers highlighted the challenges in the management of patients with multimorbidity, particularly when they alternate between traditional and/or spiritual practices, and biomedical approaches.

> “*So many times I have encountered patients at the ED (emergency department) who are known hypertensive and diabetic who stopped taking their medications on religious basis…they end up with CKD*” [Specialist 2 Tanzania].

> “*I feel like patients do not like taking lifelong medications, so they resort to herbal medication to get treated”* [General medical doctor 6 Malawi].

#### Disease severity at hospital presentation

In both Malawi and Tanzania, healthcare workers identified acuity of clinical presentation and secondary complications as key factors that influence their clinical decisions in the management of patients with multimorbidity. Less emphasis was placed in medium and longer-term management to prevent complications.

> “*The decision on how I manage patients with multiple chronic conditions is based on the clinical presentation on the day of admission and what is the most eminent condition that will kill the patient quicker*” [clinical officer 2 Malawi]

> “*In clinical management, it depends on what stage the patient presents in the hospital. Most of these chronic conditions like diabetes and hypertension have complications that we address when managing patients*” [Specialist 1 Tanzania]

### Workforce domain

Multidisciplinary team working

#### Delays in clinical decision-making

Healthcare workers had a positive attitude towards interdisciplinary team involvement in the management of patients. However, some healthcare workers expressed their frustrations over delays in clinical decision-making for managing complex patients with multimorbidity, attributing these delays to reliance on external consultations in tertiary hospitals.

> “*The challenge with the management of multimorbidity is consultations with several clinicians and the timing of their arrival. getting all specialists to review the patient takes time and it’s frustrating when the patient needs urgent care*” [Specialist 4 Tanzania]

> “*If the patient has multimorbidity and has a surgical condition, it means we need to involve the medical team and the surgical team. Getting hold of these teams is quite hard. It means their treatment may delay*” [Clinical officer 6 Malawi].

#### Healthcare worker resource and capacity

Healthcare workers highlighted challenges related to inadequate staffing, particularly a shortage of clinicians, in healthcare facilities across both Malawi and Tanzania. Urgent consultations for patients with acute decompensated complications of multimorbidity resulted in de-prioritization of non-acute patients managed in outpatient settings:

> “*On the clinical side, the clinicians are really understaffed, we only have two clinicians who are supposed to cover the ward and the clinic. we get overwhelmed by seeing many patients sometimes*” [Clinical officer 5 Malawi].

> “*We have a serious shortage of healthcare staff here, I am the only specialist, sometimes I see 40 to 50 patients a day. I think we need more manpower to handle the rising number of patients.* ” [Specialist 6 Tanzania].

Some of the healthcare workers recommended task shifting as one of the solutions to address the challenges related to the management of stable chronic patients.

> “*We need to empower nurses with protocols that they can easily follow to manage patients with hypertension and diabetes attending NCDs (non-communicable disease) clinics as with HIV clinics and care. That way we can concentrate on the acute patients and ease the burden* ” [General medical doctor 8 Malawi]

### Continuous professional development

#### Lack of post-graduate training to support multimorbidity management

Whilst multimorbidity was alluded to in some existing training materials available to clinicians in these contexts, these were not comprehensive and did not adequately address the management complexities for patients with interacting chronic diseases.

> “*I attended a training on the management of patients with HIV that discussed complications including CKD……The rest you have to read on your own how to manage multimorbidity*” [Clinical officer 4 Malawi]

> “*There are basic life support courses that I have attended to manage trauma and critical patients that touched base on the management of multimorbidity. However, they were not specific to patients with multimorbidity only*” [Specialist 2 Tanzania].

### Education and workforce planning

#### Absence of integrated management training

Healthcare workers in Malawi and Tanzania described their clinical training as focused on single diseases, limiting their ability to effectively make decisions for the management of patients with multimorbidity. To address this challenge healthcare workers suggested re-adjustments of the clinical training curriculum to address integrated disease management.

> “*There is a gap in our clinical training. We learn about disease conditions like hypertension, diabetes and CKD in isolation. When we come to patients, these conditions co-occur. We need to look at our training*” [General medical doctor 3].

> “*We didn’t have a module addressing integrated management of multimorbidity during our clinical training. We would talk about diabetes, HIV or kidney disease as separate entities* ” [General medical doctor 5 Malawi].

### Workforce disease-burden mismatch

#### Limited management at primary care and district-level facility level (unequal workforce distribution)

Clinicians in secondary care facilities perceived that patients with multiple chronic conditions were not adequately managed in lower-level facilities. They felt that this contributed to the presence of complex multimorbidity cases in hospitals, which are particularly challenging to manage. This highlights the need for improved linkage and coordination of care between facilities in these settings.

> “*Healthcare workers in lower-level facilities are the first contact of patients to the healthcare system. they have little knowledge about chronic disease management. That’s where most of the misdiagnosis and mismanagement happens* ” [General medical doctor 3 Tanzania]

> “*Healthcare workers at health centres lack adequate knowledge to manage chronic conditions effectively…. For example, a patient presents at the hospital with a stroke, but their health passport reveals they had multiple visits to different facilities where they were repeatedly mismanaged*” …. [General medical doctor 6 Malawi].

To address the linkage issues healthcare workers suggested capacity building of healthcare workers in lower-level facilities as one of the solutions.

> “*I would suggest training clinicians in dispensaries and health centres on how to manage patients with NCDs, because this is where the majority of the patients referred to tertiary hospitals come from* ” [Specialist 1 Tanzania]

> “*There is no mentorship and supervision of health centres to see how they manage patients, including how they choose to start medications for patients like those with hypertension how to adjust the doses. ”* [Clinical officer 1 Malawi]

### Service delivery domain

#### Treatment interactions

##### Lack of guidelines for multimorbidity care

The perceived lack of clinical practice guidelines specific to multimorbidity is a barrier to standardised and effective clinical management for multimorbidity. Healthcare workers highlighted the challenge of balancing treatment and drug interactions during the management of patients with multimorbidity.

> “*Most of the guidelines are single disease based, like we have the HIV guideline, TB guideline and the diabetes mellitus guideline and they are different. The clinician needs to integrate them when they have a patient which is very challenging…and you need to account for drug interactions*” [Specialist 4 Tanzania].

> “*We have the standard treatment guideline (the blue book), and we have protocols for some chronic conditions. But we don’t have something specific on multimorbidity in our setting… There are some grey areas because it’s not clear for these patients what we should and should not give*” [General medical doctor 5 Malawi].

To address treatment interaction, some of the clinicians reported the use of digital applications, formulary books (including the British National Formulary [BNF]) and consultations with pharmacists.

> “*I use my phone and apps like Medscape to check for drug interactions when prescribing to patients. Sometimes I consult the pharmacist or google the internet to make sure I can or cannot prescribe a certain drug*” [General medical doctor 1 Tanzania].

> “*I usually use the BNF (British National Formulary) to check whether the drugs I prescribed interact. But there are some of the drugs you know through experience that they will interact like the TB and HIV drugs* ” [General medical doctor 8 Malawi].

#### Service availability and access

##### Limited investigations and availability of medicines

The differences in healthcare resource allocation between tertiary and district hospitals resulted in limited availability of diagnostic tools and medicines in district hospitals. Healthcare workers in district hospitals in Malawi and Tanzania reported occasionally directing patients to private clinics for prescriptions or referring them to tertiary hospitals for further investigations and treatment.

> “*The challenges with the management of patients with multiple chronic conditions is stock-outs of medicines. For, instance it’s been a while since we had the second line of antihypertensives. Sometimes we don’t have even the first line and patients miss their drugs.*….. “[Nurse 15 Malawi].

> “*In peripheral facilities, the diagnosis and management of multimorbidity is challenging because of limited resources. Here in Muhimbili [a tertiary hospital] if I suspect diabetes, I can confirm the diagnosis with HbA1c, but I cannot do that in the dispensary in Lindi (primary care) ”* [General medical doctor 3 Tanzania]

The limited availability of diagnostics and clinical expertise significantly influenced healthcare workers decisions in the management of multimorbidity, leading to unnecessary referrals to higher-level centres.

> “*Sometimes the referrals are due to lack of investigations and expertise to interpret the investigations in lower-level facilities”* [Specialist 3 Tanzania].

> “*The electrolytes reagents in our lab, most of the time, are out of stock, so we send our patients to Nguludi (private clinic) and sometimes we contact Queens Elizabeth Central Hospital and refer the patient for further investigations and treatment* “[Clinical officer 3 Malawi].

#### Policies to integrate care across organizations and sectors

##### Lack of integrated disease management policies

Healthcare workers in Malawi and Tanzania reported absence of integrated policies to guide the management of patients with multimorbidity. Concerns were raised about inadequate funding and investment in the management of NCDs compared to the investment in HIV care leading to a lack of continuity of care for patients across healthcare systems.

> “*With HIV services, we are doing so well because we have a lot of support. We have individuals and organizations that have invested so much in HIV care. but in the case of hypertension and diabetes, we still have a long way to go. One thing we struggle the most with is awareness.* ” [General medical doctor 8 Malawi].

> “With *HIV there is a massive investment on prevention and awareness in the community and awareness is big compared to the awareness on NCDs like hypertension and diabetes*” [Specialist 1 Tanzania].

### Financing domain

#### Out-of-pocket costs

##### Healthcare access and treatment cost

Healthcare workers noted that patient access to healthcare and treatment costs significantly influenced their decisions in managing multimorbidity. In Tanzania, where public healthcare is not free to patients, healthcare workers face challenges in balancing quality care for patients without insurance who face potentially catastrophic out-of-pocket expenditures.

> “*Most of the patients with multimorbidity are poor and do not have health insurance, they pay cash. If I write two or three investigations and prescribe medications afterwards, mostly cannot afford and that’s a challenge to move on with the management* “[Specialist 4 Tanzania]

> “*For instance, the patient has hypertension, diabetes and renal failure, does need dialysis which costs about $50 per week and has no health insurance. I know the majority cannot afford this cost…. yes, there are exemptions but there is a cap to that too they need to pay* ” [Nurse 3 Tanzania].

However, for patients with multimorbidity who had health insurance, clinical decision-making was described as less challenging, as it allowed for a broader range of diagnostic and treatment options to be explored.

> “*For patients who have CHF and NIHF, there are a lot of options for investigations and medicines. The challenge is with multimorbidity patients who pay cash [uninsured], most of the time they don’t have the money. ”* [General medical doctor 4 Tanzania]

In Malawi where public healthcare is free, the lack of money for transportation was ascribed to missed appointments and advanced disease presentation.

> “*Some patients miss their clinic appointment like in the HIV clinic because they have to travel a long distance, and they have no money for transport. MSF is supporting some of the patients too with transport cash*” [Nurse 14 Malawi].

## Discussion

We found that clinical decision-making for the management of patients with multimorbidity is influenced by a combination of patient-level, healthcare provider, and health system factors. Patient-specific factors such as the severity of diseases; chronic condition health literacy; and socioeconomic status significantly influenced clinical decisions among healthcare workers in Malawi and Tanzania. Healthcare worker shortages and limited integrated chronic disease training, including insufficient consideration of multimorbidity within clinical practice guidelines, also impact clinical care delivery. High out-of-pocket costs for chronic conditions also appear to influence clinical decision-making among healthcare workers. Our findings emphasize the need to strengthen integrated care; expand workforce capacity; update clinical guidelines; and strengthen healthcare financing through the expansion of universal health coverage (and insurance programmes where applicable) to address the growing multimorbidity burden in SSA.

We observed that secondary healthcare workers prioritised management of patients with acute presentations in both Malawi and Tanzania, in alignment with other studies (43). This approach often fails to address broader risk factors and social determinants of health, missing opportunities to prevent further disease progression. To achieve integrated holistic patient care, health systems could change from reactive to preventative patient-centred healthcare delivery (44, 45). Other studies advocate for healthcare workers to expand their approach to disease management, integrating prevention and health promotion alongside acute hospital care (46). However, addressing barriers such as resource limitations, staff shortages, and high workloads is essential for successful adoption of proactive chronic care in hospitals in Malawi and Tanzania.

Healthcare workers noted that the lack of integrated chronic disease management clinical training and practice guidelines impacts on effective decision-making for multimorbidity care. Similar observations were noted among general practitioners and physicians involved in the management of patients with multimorbidity in high income countries (15, 47, 48). Vertical programs with condition-specific funding (49); a high burden of infectious diseases such as malaria (12); and limited focus on multimorbidity prevention contribute to the curative bias in sub-Saharan Africa (9). Throughout this study, participants reported engagement with continuous professional development (CPD), however, these activities were not specific to multimorbidity. CPDs have been shown to play a crucial role in empowering healthcare workers with updated knowledge, skills and competences in LMICs (50, 51). Further, a report from the WHO suggests that CPD investment for chronic diseases management strengthens healthcare systems, reduces preventable complications, and enhances the quality of patient-cantered care (52).

Healthcare workers described insufficient recognition and management of patients with multimorbidity occurs in lower-level facilities in Malawi and Tanzania. This was ascribed to inadequate clinical mentorship and training. These perceptions may reflect a disconnect between primary and secondary care delivery within the health system, a potentially important nexus of intervention. Previous work demonstrates that enhanced collaboration between primary and secondary care providers is associated with improved patient outcomes and reduce unnecessary healthcare costs for multimorbidity management (53). Healthcare workers suggested that increased mentorship for healthcare workers in lower-level facilities and task shifting could be helpful to achieve improvements. A scoping review on kidney and cardiovascular disease care in Africa found that task shifting enhanced access and efficiency in chronic disease management (54). Although task shifting may improve efficiency and ease the burden on higher-level facilities, structured implementation that addresses potentially conflicting healthcare delivery priorities for staff in existing lower-level facilities with clear guidance is crucial.

The lack of specific clinical guidelines for the management of multimorbidity in Malawi and Tanzania presents a significant challenge for healthcare workers in making evidence-based clinical decisions. Without guidelines, healthcare workers rely on individual experiences, or single-disease guidelines (47), which may not address the complexities of the management of multimorbidity. The challenges of multimorbidity guidelines in Malawi and Tanzania mirror those in other LMICs, where comprehensive guidelines and integrated care models remain undeveloped or unimplemented (55, 56). In this study, healthcare workers noted that the lack of clinical guidelines complicates multimorbidity management. The absence of standard guidelines for multimorbidity management has been linked to treatment interactions, increased risks of adverse events, polypharmacy, and inconsistent treatment approaches (57, 58), implementation of local and internationally developed (e.g. WHO package of essential noncommunicable (PEN) disease interventions (59)), context-relevant integrated care guidelines, building on existing work, could substantially improve multimorbidity management and reduce disparities in care.

The proper management of multimorbidity requires consistent access to diagnostics, medications and monitoring (60), with healthcare financing shaping access (61). Healthcare workers in Tanzania cited high out-of-pocket costs for uninsured patients, while those in Malawi highlighted transport costs as a barrier despite free public healthcare. A scoping review similarly found that high out-of-pocket costs drive cost-related non-adherence, forcing patients with multimorbidity to prioritize treatments, resulting in suboptimal care and poorer health outcomes (62). This highlights the need for sustainable healthcare financing in SSA, where out-of-pocket payments remain the dominant funding model (63). Recurrent stock-outs force healthcare providers to prescribe less effective or more expensive alternatives, increase household financial strain and risk to catastrophic health expenditure and worsening of treatment adherence (16, 64).

The key strength of this study is the representation of multiple healthcare worker cadres including doctors, clinical officers, nurses and pharmacists from district and tertiary hospitals in Malawi and Tanzania, who provided broad, generalisable perspectives on multimorbidity management to similar settings. This representation of different healthcare worker cadres from both contexts enhanced the understanding of multimorbidity management in Malawi and Tanzania and highlighted the existing care delivery gaps. Whilst recent policy reforms in Tanzania have increased the prominence of healthcare insurance, the Tanzanian and Malawian healthcare systems share similarities, making these findings relevant to other resource-limited countries in sSA. We note, however, several limitations to our study. These include a limited number of healthcare workers in certain categories, which may not fully represent the experiences of all participants. Another limitation of this study is the exclusion of primary level healthcare workers, who are the first point of healthcare contact for the majority of patients in the two countries. Moreover, the findings of this study are based on self-reported experiences of healthcare workers, which may be subject to social desirability bias. Priority setting partnerships (e.g. using the James Lind Alliance methodology (65)) bringing together key stakeholders, inclusive of patient perspectives would be a useful next step to set the agenda for multimorbidity management in this context.

This study demonstrates that a combination of patient, healthcare workers and health system factors influence healthcare worker clinical decisions for the management of multimorbidity. The most prominent factor is the lack of clinical training on integrated management of chronic diseases among healthcare workers in Malawi and Tanzania. For healthcare workers to provide the best integrated management care for patients with multimorbidity, we recommend re-orientation of the clinical training curricula by policy makers and clinical training institutions. Furthermore, our study highlights that the absence of multimorbidity clinical practice guidelines impacts healthcare workers clinical decisions for multimorbidity management. These findings provide key insights into factors influencing clinical decisions for the management of patients with multimorbidity in district and tertiary hospitals in Malawi and Tanzania. The insights may help to close the gap in knowledge and practice to improve clinical outcomes of patients with multimorbidity in sSA.

## Data Availability

The data supporting the findings of this study are available from the corresponsing author [IGS] upon request.

## Author contributions

Study conceptualization: Ibrahim G. Simiyu, Charity Salima, Juma Mfinanga, Adamson Muula, Matthew P Rubach, Blandina T Mmbaga, Sarah Urasa, Miriam Taegtmeyer, Francis Sakita, Eve Worrall, Hendry R. Sawe, Paul Dark, Felix Limbani, Ben Morton

Data curation: Ibrahim G. Simiyu, Gift T Banda, Nateiya Mmeta Yongolo, Grasiana Kimario, Martha Oshosen, Gimbo Hyuha,

Formal analysis; Ibrahim G. Simiyu, Hendry R. Sawe, Paul Dark, Felix Limbani, Ben Morton

Funding: Ben Morton, Hendry R. Sawe, Juma Mfinanga, Matthew P. Rubach, Adamson Muula, Paul Dark, Miriam Taegtmeyer, Blandina T Mmbaga, Felix Limbani, Jammie Rylance, Eve Worrall, Ben Morton.

Investigation: Ibrahim G Simiyu, Gift T Banda, Nateiya M Yongolo, Grasiana Kimario, Martha Oshosen, Gimbo Hyuha

Methodology: Ibrahim G Simiyu, Hendry R. Sawe, Paul Dark, Felix Limbani, Ben Morton

Project Administration: Ibrahim G Simiyu, Gift T Banda, Nateiya M Yongolo, Sangwani Salimu, Grasiana Kimario, Marth Oshosen, Jacob Phulusa, Gimbo Hyuha, Juma Mfinanga, Rhona Mijumbi, Hendry R Sawe, Matthew P Rubach, Jammie Rylance, Felix Limbani, Eve Worrall, Ben Morton

Supervision: Hendry R Sawe, Juma Mfinanga, Sarah Urasa, Paul Dark, Jammie Rylance, Eve Worrall, Felix Limbani, Ben Morton.

Validation: Ibrahim G Simiyu, Gift T Banda, Hendry R Sawe, Paul Dark, Felix Limbani, Ben Morton

Writing-original draft: Ibrahim G Simiyu, Hendry R Sawe, Paul Dark, Felix Limbani, Ben Morton

Writing-review & editing: Ibrahim G Simiyu, Gift T Banda, Nateiya M Yongolo, Sangwani Salimu, Hendry R Sawe, Paul Dark, Felix Limbani, Ben Morton.

## Acknowledgement

Sanjura Biswalo, Yesse Bumija, Robert Chuwa, Rose Freddy, Mwamini Kacheuka, Frank Kimaro, Zanuni Kweka, Rachel Mangoni, Philoteus Sakasaka, Constantine Tarimo, Gidion Tesha, Safina Baleche, Yusuph Chimpaye, Frank Gugu, Naftari Mahimbo, Ramadhani Mashoka, Vicky Mlele, Hussein Abdallah Moremi, Noela Mpili, Herieth Cliff Mushi, Nsajigwa Mwakyambiki, Benjamin Paulo Mwenda, Abdulaziz Abdallah Nassoro, Nuhu Richard, Chiku Simbano, Marlen Chawani, Augustine Choko, Marlen Chewani, Beatrice Chinoko, Sylvester Kaimba, Maureen Kandiero, Lucy Keyala, Florence Malowa, Peter Mandala, Mercy Mkandawire, Matthew Mlongoti, Bright Mnesa, Albert Mukatipa, Alfred Muyaya, Deborah Nyirenda, Diana Msindira, Genesis Chowe, Yusuf Iqbal, Joanna Jozefiak, Nicola Desmond, Firdaus Hafidz, Amy Smith.

## List of abbreviations

CPD: Continuous Professional Development
CKD: Chronic Kidney Disease
CHF: Community Health Fund
NHIF: National Health Insurance Fund
MSF: Médecins Sans Frontières

